# Patient-specific *in silico* prediction of outcomes of partial continuous-flow LVAD treatment in peripartum cardiomyopathy

**DOI:** 10.1101/2024.09.18.24313803

**Authors:** Juliet Nagawa, Kevin L. Sack, N. John Nchejane, Y. Dodzi Motchon, Mazin S. Sirry, Sarah Kraus, Neil H. Davies, Ntobeko A.B. Ntusi, Thomas Franz

## Abstract

Patients with severe peripartum cardiomyopathy (PPCM) often receive mechanical circulatory support with good outcomes. However, mechanisms underlying the functional improvements are poorly understood for patients with different PPCM characteristics. This study investigated effects of partial, continuous-flow left ventricular assist device (LVAD) support on cardiac function and mechanics in patients with different PPCM severity. Patient-specific biventricular finite element models of six patients with different PPCM severity were developed from magnetic resonance images and combined with a circulatory system model, including variable LVAD support. Ventricular function and myocardial mechanics were predicted and changes due to LVAD support were quantified. The LVAD support decreased LV myofiber stress and increased ejection fraction (EF). EF increased steadily (two patients), fluctuated (two patients), or peaked before a steady decrease (two patients) with increasing LVAD speed. Improvement due to LVAD support was greater for PPCM patients with higher disease severity than those with lower disease severity. The LVAD and native LV jointly generated stroke volume (SV) in four patients, and the LV contribution diminished with increasing LVAD speed. In the two patients with the lowest EF, the LVAD was the sole source of SV. The improvement of cardiac function and mechanics due to LVAD support in PPCM exceeds that reported for chronic heart failure due to ischemia. However, the predicted variability of the LVAD benefits with PPCM severity and mechanical support level suggests the need and potential for further studies to guide clinicians in selecting personalised treatment parameters required for optimised LVAD therapy for each PPCM patient.

## 1. Introduction

Cardiomyopathies ranked as the second leading cause of heart failure in Sub-Saharan Africa, with Southern Africa exhibiting the highest prevalence (40%) of cardiomyopathies, according to Agbor [1]. Moreover, peripartum cardiomyopathy (PPCM) was among the most common cardiomyopathies in Sub-Saharan Africa [1]. In South Africa, PPCM is the largest contributor to maternal death [2, 3]. In addition, studies have shown that women of African ethnicity have a higher risk of developing PPCM and obtaining poorer outcomes than women of other ethnicities [4, 5]. Moreover, poorer maternal and neonatal outcomes often present in African countries with poor socioeconomic characteristics [6].

Therapy efficacy for PPCM has mixed outcomes. While many patients respond positively to treatment, some never recover, and others die despite optimal therapy [7, 8]. PPCM is associated with high recovery rates compared to other cardiomyopathies - about 59% of PPCM patients have recovery of LV function, and one in 10 PPCM patients dies within the first year of diagnosis [8, 9]. In cases of severe PPCM, the left ventricular assist devices (LVAD) have been used among other mechanical circulatory support therapies such as the biventricular assist device, peripheral veno-arterial extracorporeal membrane oxygenation pump, and the total artificial heart [10].

These therapies have achieved remarkable left ventricular (LV) recovery and high survival rates for PPCM patients [11, 12]. For instance, Berliner [11] reported that LVAD treatment resulted in a high recovery rate of 25% with successful LVAD explantation and a high survival rate of 91.7%. Djordjevic [13] reported a ventricular recovery rate of 20% and a survival rate of 87%. However, the mechanisms associated with the reported positive outcomes of device therapy in PPCM are poorly understood and it is not yet clear how patient-specific characteristics influence outcomes of the LVAD support.

Due to their high costs, circulatory support devices, including LVADs, have been administered primarily in developed countries. In contrast, their use has been limited in Africa [2] despite generally higher PPCM incidence (e.g. Nigeria 98:10,000 live births [14], South Africa 10:10,000 [15]) than in developed countries (e.g. Germany 6.7:10,000 [17], Japan 0.5:10,000 [18]) except the USA with a high proportion of people of African ethnicity (11.8:10,000 [16]). Moreover, causes and outcomes of heart failure in Africa are different compared to high-income countries; African patients present at a younger age and exhibit higher mortality rates [19, 20]. It is therefore important to investigate outcomes of LVAD treatment for African patients.

The Heart Failure Society of South Africa recently developed guidelines for the effective use of LVADs in low-resource settings [21]. Rapid technological advancements and increasing emphasis on affordable healthcare solutions are expected to decrease the costs of healthcare technology in the near future. In this context, a better understanding of the therapeutic effects of LVAD support in PPCM can lead to considerable improvements in the treatment of these patients and be adapted for other heart failure patients in Africa.

*In silico* studies can contribute to gaining knowledge on cardiac mechanics and the effects of circulatory support devices for PPCM patients with different disease severities. Finite element models have been used to investigate cardiac disease and therapeutic mechanisms for heart failure. These models enable the modification and analysis of therapeutic and disease parameters that may not be possible in experimental studies due to ethical considerations. For example, such studies have provided insights into biomechanical mechanisms of heart failure and myocardial infarction and the development and efficacy evaluation of therapies involving LVADs [22, 23], biomaterial injections [24, 25], valve repair [26] and surgical restoration [27].

In addition, the rapid increase in demand for personalised medicine and the advancement of technology have encouraged the use of patient-specific computation models to quantify the effects of heart failure therapies on cardiac function, mechanics and remodelling during heart failure, which is not easily achieved with other methods [28].

The current study aimed to investigate how disease severity affects the efficacy of partial LVAD support in South African PPCM patients using patient-specific finite element models.

## 2. Methods

### 2.1. Patient data

The definition of PPCM by Sliwa [29] was used as an inclusion criterion, and patients who did not fit this definition were not considered in the current study. Six female patients with PPCM, with an age range from 19 to 41 years, were included in the current study.

*In vivo* cardiac magnetic resonance imaging (cMRI) and echocardiographic data for six adult PPCM patients were obtained from existing records at Groote Schuur Hospital. A longitudinal dataset was used, and information on the patient’s diagnosis at baseline and recovery status after treatment was available from the clinical records at the beginning of this study. Recovery implied that the patient’s cardiac function normalised after treatment, whereas non-recovery implied that the patient’s cardiac function did not normalise after treatment. A pseudonym was used for each patient representing LV ejection fraction (EF) after diagnosis at baseline, recovery status (R: recovered, N: not recovered), and survival status (A: alive, D: deceased) from clinical records at the beginning of the current study: R45A, N39A, R20A, N18A, N12A and N06D. Four patients had severely abnormal LV function with EF < 30%, based on the recommendations of the American Society of Echocardiography and the European Association of Cardiovascular Imaging [30, 31]. One patient (N39A) had LV concentric remodelling (normal LV mass ≤ 95 g/m^2^ with increased relative wall thickness > 0.42), whereas the other five patients had normal LV geometries as defined by Lang [30] (Table 1).

**Table 1:**
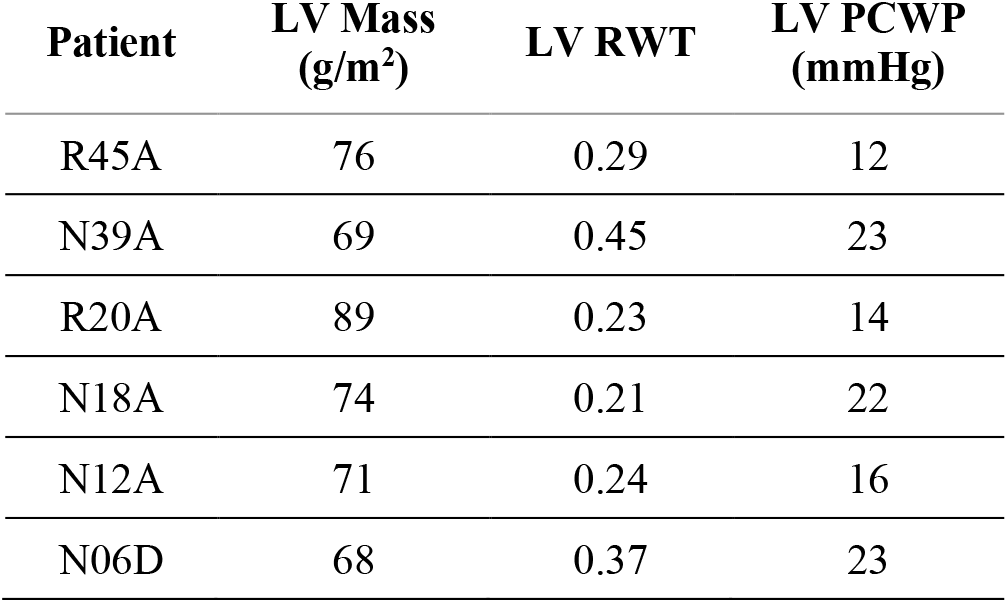
Clinical *in vivo* cardiac metrics for the left (LV) and right ventricle (RV) of PPCM patients (PCWP: Pulmonary capillary wedge pressure, RWT: Relative wall thickness).

| Patient | LV Mass<br>(g/m<sup>2</sup>) | LV RWT | LV PCWP<br>(mmHg) |
| --- | --- | --- | --- |
| R45A | 76 | 0.29 | 12 |
| N39A | 69 | 0.45 | 23 |
| R20A | 89 | 0.23 | 14 |
| N18A | 74 | 0.21 | 22 |
| N12A | 71 | 0.24 | 16 |
| N06D | 68 | 0.37 | 23 |

### 2.2. Development of cardiac geometries

Three-dimensional biventricular cardiac geometries truncated at the base were reconstructed from short-axis cMRI data at early diastole to approximate the unloaded *in vivo* configuration (Simpleware ScanIP, Synopsys, Mountain View, CA, USA). The geometries were meshed using approximately 85,000 quadratic tetrahedral elements (Figure 1 a), i.e. 85,078 for model R45A, 85,656 for N39A, 85,075 for R20A, 85,321 for N18A, 85,040 for N12A, and 85,026 for N06D, following a mesh convergence study. The mesh sizes allowed adequate element distribution across the ventricular wall thickness, essential for accurate fibre representation and determination of stress and strain. Similar findings were reported in a mesh convergence study by Sack [32].

**Figure 1:**
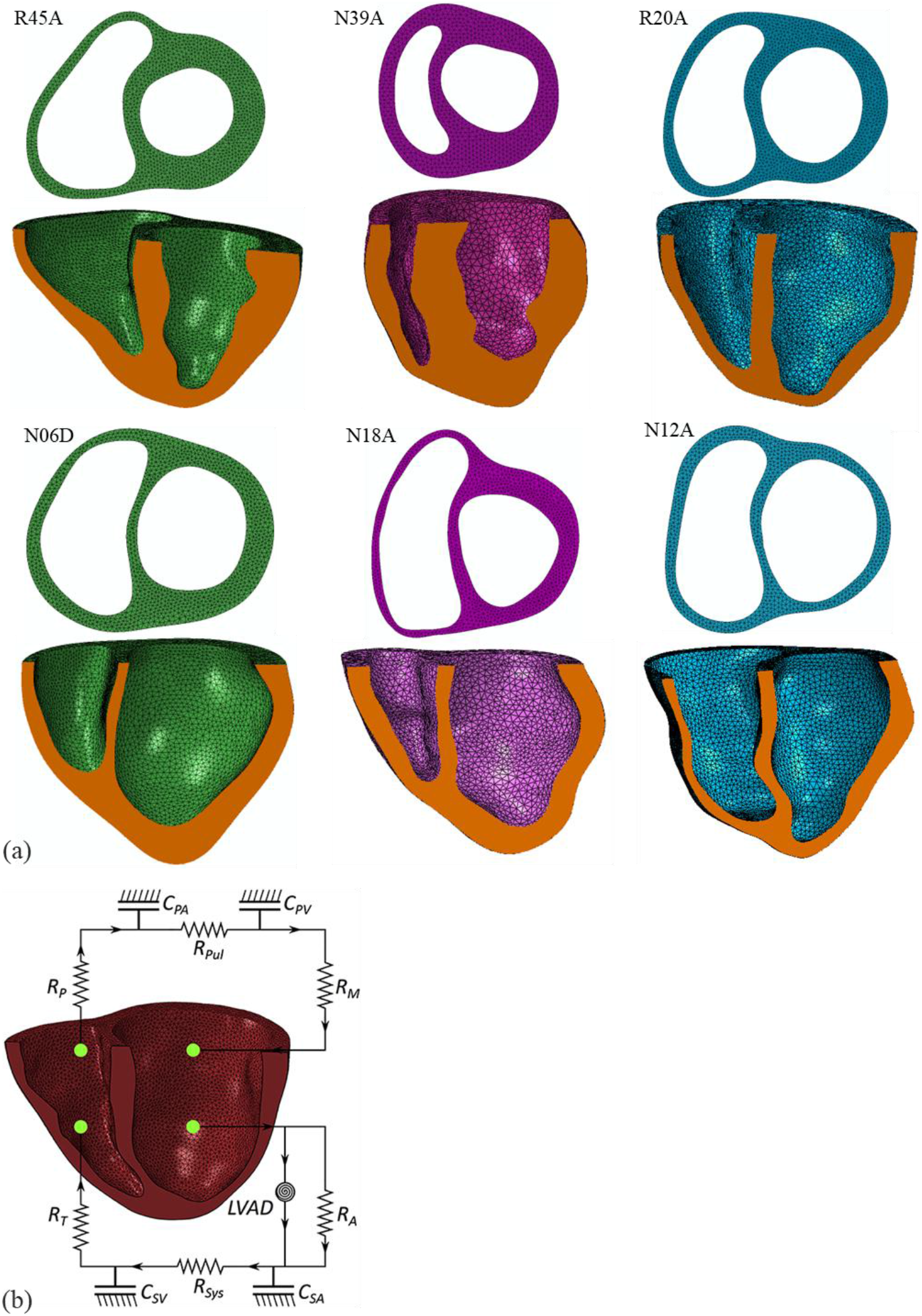
a) Short- and long-axis views of biventricular meshed geometries of six PPCM patients illustrating the wall thickness of the left and right ventricular free wall and septal wall. In each image, the LV is on the right, and the RV is on the left. Ejection fraction is indicated by the number in the patient code. b) Schematic of coupling of biventricular geometry with circulatory system and LVAD [22]. R_M_: mitral valve resistance, R_A_: aortic valve resistance, C_SA_: systemic arterial compliance, R_SYS_: systemic arterial resistance, C_SV_: systemic venous compliance, R_T:_ tricuspid valve resistance, R_P_: pulmonary valve resistance, C_PA_: pulmonary arterial compliance, R_P_: pulmonary valve resistance, C_PV_: pulmonary venous compliance, LVAD: left ventricular assist device.

### 2.3. Development of cardiac finite element models

The meshed biventricular cardiac geometries were imported into Abaqus CAE (Dassault Systèmes, Providence, USA).

#### 2.3.1. Boundary conditions

The LV and RV cavities were closed with caps created using two-dimensional triangular surface elements between basal endocardial nodes and central cavity nodes (one for the LV and three for the RV). The cavity caps facilitated the calculation of cavity volumes to assess ventricular function. The endocardial and central nodes at the base were fixed in the cardiac longitudinal direction to prevent rigid body motion of the biventricular geometry. The degrees of freedom of the central nodes were coupled to the average motion of the endocardial nodes, allowing movement of the endocardial and epicardial nodes at the base in the short-axis plane.

#### 2.3.2. Myofibre orientation

Using a rule-based method, the myofibre orientation was implemented as a linear variation of the helix angle from −60º on the epicardium to +60º on the endocardium in all elements, except those forming the cavity caps, using a custom MATLAB script (The MathWorks, Inc., Natick, Massachusetts, United States) [22]. The LV and RV free walls and the septal wall had the same myofiber description [22].

#### 2.3.3. Constitutive law for myocardium

The description of the material properties of the myocardium was based on Sack [32]. The passive material properties were defined with an anisotropic hyper-elastic formulation [33]. The strain energy potentials *Ψ*_iso_ and *Ψ*_vol_ govern the isochoric and volumetric responses, respectively, and capture the experimentally observed exponential and anisotropic elastic response and near-compressibility [34]:

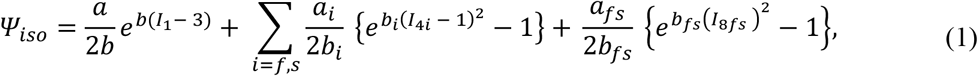

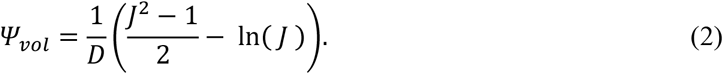

All parameters are described in Table 2. The active material law is based on a time-varying elastance model by Guccione [35], and a more detailed description can be found in Sack [22]. The active tension *T*_*a*_ is:

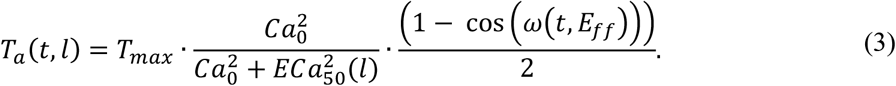

**Table 2:** Constitutive parameters for the passive and active material response of the myocardium [32].

| Parameter | Description |
| --- | --- |
| $a, b$ | Scales isotropic tissue response |
| $a_f, b_f$ | Scales additional stiffness in fibre direction |
| $a_s, b_s$ | Scales additional stiffness in sheet direction |
| $a_{fs}, b_{fs}$ | Scales coupling stiffness in fibre and sheet directions |
| $A_i$ | Vector in direction $i$ |
| $I_1$ | First deviatoric strain invariant |
| $I_{4i}$ | Pseudo-invariant defined as $A_i \cdot C \cdot A_i$ |
| $I_{8fs}$ | Pseudo-invariant defined as $A_f \cdot C \cdot A_s$ |
| $C$ | Right Cauchy-Green tensor |
| $D$ | Multiple of bulk modulus ($K = 2/D$) |
| $J$ | Third deformation gradient invariant |
| $t_0$ | Time to reach peak tension |
| $m$ | Governs slope of relaxation |
| $b$ | Governs length of relaxation |
| $l_0$ | Sarcomere length below which no active force develops |
| $B$ | Governs shape of peak isometric tension-sarcomere length relation |
| $Ca_0$ | Peak intercellular calcium concentration |
| $Ca_{max}$ | Maximum intercellular calcium concentration |
| $T_{max}$ | Contractility scaling factor (value directly scales ejection fraction) |
| $E_{ff}$ | Lagrangian tensor component aligned with local muscle fiber direction |
| $n_s$ | Fraction of active tension to be applied in sheet direction |
| $n_n$ | Fraction of active tension to be applied in sheet-normal direction |
| $l_r$ | Initial sarcomere length |

The maximum active tension *T*_*max*_ is multiplied by two terms governing the calcium concentration and the time of contraction, respectively. The formulation of the active tension considers sarcomere length *l*, which affects the stress components in the fibre direction and captures the Frank-Starling effect [36]. The total stress in the fibre direction, σ_*f*_, and sheet direction, σ_*s*_, are:

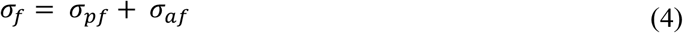

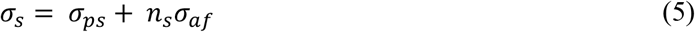

where σ_*pf*_ is the passive stress in fibre direction, σ_*af*_ is the active stress in fibre direction, and σ_*ps*_ is the passive stress in sheet direction. A fraction of the active stress in fibre direction that is transferred to the sheet direction was accounted for by a scalar *n*_*s*_ *ϵ* (0,1) [22, 32].

### 2.4. Patient-specific optimisation of myocardial constitutive parameters

#### 2.4.1. Passive mechanical properties

To obtain patient-specific passive material parameters, the linear (*a*_*i*_) and exponential (*b*_*i*_) terms in Eqn. (1) were optimised by minimising the error between the LV end-diastolic pressure values predicted by the FE model and those obtained from the patient’s Klotz curve [37]. The LV pressure and volume values at the end-diastolic time point were used in the method described by Klotz [37] to estimate each patient’s LV end-diastolic pressure-volume relationship (Klotz curve). The LV end-diastolic volume was calculated using the ejection fraction from clinical records, and the volume V_0_ was determined from the segmented cardiac geometry. Since the LV end-diastolic pressure was unavailable, it was assumed to be equal to the pulmonary capillary wedge pressure (PCWP, Table 1) computed for each patient using Nagueh’s formula [38]:

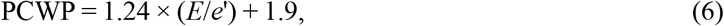

where *E* is the mitral inflow velocity and *e*’ is the average of the septal and lateral annular velocities from clinical records.

The sequential least-squares algorithm was used to minimise the error between the LV end-diastolic pressure predicted with the FE model and estimated from the patient’s Klotz curve:

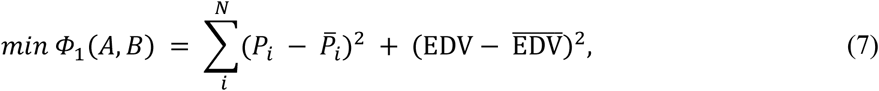

where *N* is the total number of points on the pressure-volume curve, *P*_*i*_ and EDV are pressure and end-diastolic volume predicted with the FE models, 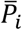 is the pressure derived from the patient’s Klotz curve, and 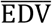 is calculated using the ejection fraction from clinical records and the volume of the unloaded left ventricle, V_0_, determined from the segmented cardiac geometry. A scalar (*A*) and an exponential multiplier (*B*) were used to uniformly scale the linear (*a*_*i*_) and exponential (*b*_*i*_) terms, respectively, in Eqn. (1). All PPCM models were assigned identical initial parameter values for healthy myocardium calibrated by Sack [32] using experimental tri-shear data from Sommer [39]: a = 1.05 kPa, b = 7.542, a_f_ = 3.465 kPa, b_f_ = 14.472, a_s_ = 0.481 kPa, b_s_ = 12.548, a_fs_ = 0.283 kPa, and b_fs_ = 3.088. The mean squared error of the LV pressure of the Klotz method and the *in silico* prediction was used to assess the quality of the optimisation.

#### 2.4.2. Active mechanical properties

The contractility scaling factor *T*_*max*_ was optimised so that the models produced clinically recorded stroke volumes. The initial value for all models was *T*_*max*_ = 100 kPa. The LV long-axis shortening (LVLS) was included in the objective function, Eqn. (8), to ensure physiological deformation during contraction as observed in the cMRI data for each patient. The sequential least-squares optimisation algorithm in Python was used with Abaqus as a forward solver:

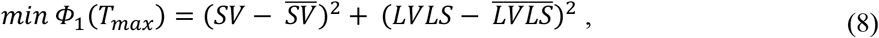

where SV and LVLS are FE predicted stroke volume and LV long-axis shortening, and 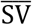 and 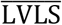 are the corresponding clinical *in vivo* values. 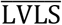 was calculated with [40]:

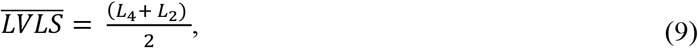

with

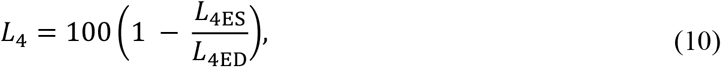

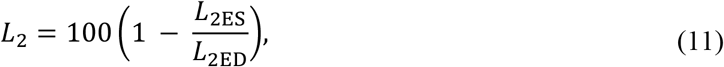

where *L*_4_ and *L*_2_ are the long-axis shortening in the 4-chamber and 2-chamber view, respectively, *L*_4ED_ and *L*_2ED_ are the lengths at end-diastole, and *L*_4ES_ and *L*_2ES_ are the lengths at end-systole. The length values were manually extracted from cine cMRI sequences of the respective chamber views.

### 2.5. Circulatory system and simulation of cardiac cycles

#### 2.5.1. Models without LVAD

A circulatory system comprising pulmonary and systemic circuits was coupled to the biventricular models as described by Sack [32]. The coupling of the lumped circulatory system and the mechanical function was carried out in Abaqus. Compliance and resistance parameters represented fluid flow between the pulmonary circuit (C_PA_ and C_PV_), heart, and systemic circuit (C_SA_ and C_SV_) (Figure 1 b).

A brief description of the volume, pressure, compliance, resistance and fluid exchange relationship within the circulatory system is provided here. Additional information can be found in the Abaqus Theory Guide [41]. The relationship between the volume *V*_*i*_, pressure *P*_*i*_ and compliance *k*_*i*_ in the fluid cavity chamber *i* is:

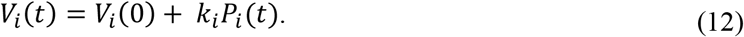

Unidirectional flow between the compliance vessels was governed by pressure gradients:

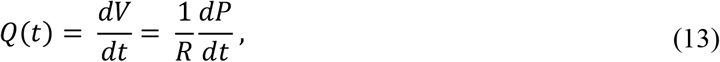

where *Q* is the flow, *V* is the volume, *P* is the pressure, and *R* is the resistance. During contraction, pressure increases in the ventricles, and flow is permitted when it exceeds the pressure in the connected outflow chamber. Volume in the circulatory system was assumed to be constant.

Each compliance vessel was loaded with fluid until the LV reached end-diastolic pressure and volume before the first cardiac cycle was initiated. The cardiac cycle comprised active contraction (480 ms) and passive filling (300 ms), corresponding to a heart rate of 77 beats per minute. Multiple cardiac cycles were simulated to facilitate solution convergence of the pressure-volume curve.

#### 2.5.2. Implementation of LVAD support

To simulate LVAD support, a secondary connection from the LV to the systemic circuit was added in the circulatory model (Figure 1 b). The flow in the secondary connection, i.e. the LVAD, was continuous and unidirectional, and introduced fluid exchange between the LV cavity and the arterial system. Only the effects of the LVAD were simulated, and fluid flow from the LV and the LVAD contributed to the arterial system.

The output from the LVAD to the arterial system was varied to simulate operational speeds of the LVAD of 8,000, 9,000, 10,000, 11,000 and 12,000 revolutions per minute (rpm). For each operational speed, the flow rate profile was coupled to the pressure difference between the inflow and outflow cavities to which the pump was connected. The profiles were based on experimental flow rates of the Heartmate II^TM^ LVAD for pressure differences varying between 0 and 200 mmHg, described by Sack [22]. Flow rates were linearly interpolated and extrapolated between discrete values to provide a continuous flow-rate representation for any pressure difference encountered by the model. Apart from the LVAD, all other fluid exchanges between cavities in the computational models were driven by pressure differences.

### 2.6. Data and statistical analysis

The patient-specific models with optimised material parameters were used to simulate multiple cardiac cycles until solution convergence was reached. Convergence was evaluated with the mean-squared error of the LV pressure and volume values from the pressure-volume loops of two subsequent cardiac cycles.

Results from end-diastole and end-systole of the 6^th^ and 10^th^ cardiac cycle were analysed for models without and with LVAD support, respectively. End-diastole and end-systole were determined from the pressure-volume curves as the time points before isovolumetric contraction and isovolumetric relaxation, respectively.

The terms’ average’ and ‘group average’ interchangeably refer to the arithmetic mean computed using values of more than one model, e.g., several patients. In contrast, the terms’ mean’ or ‘patient mean’ interchangeably refer to the arithmetic mean computed from the element values of a single model, i.e., patient.

Results from models without (untreated) and with LVAD (treated) were compared, and statistical analyses were performed for all patients.

Myofibre strains and stresses were normalised by element volume to eliminate potential effects from different element sizes when quantifying mean values. The volumetrically normalised mean value 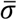 of the entire cardiac geometry was calculated using:

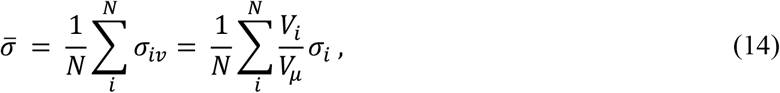

where *σ*_*iv*_ is the volume-normalised stress (or strain) of the individual element, *N* is the number of elements, *V*_*i*_ is the volume of element *i, V*_μ_ is the mean volume for all elements, and *σ*_*i*_ is the stress (or strain) value of element *i* in the model. This normalisation was only applied to the analysis of the mean value of stress and strain for each model. Contour plots illustrating the spatial distribution of stress (or strain) present the actual (not normalised) stress (or strain) values.

Considering all patients, statistical comparisons of average myofibre stress and strain, respectively, were performed between models without LVAD support and models with LVAD support at operational speeds of 8,000, 10,000 and 12,000 rpm.

Results were reported as mean ± standard deviation (SD) or as median (interquartile range). Statistical tests used include the Shapiro-Wilk and the Kolmogorov-Smirnov tests for normality. Parametric tests used included the Student t-test and Welch test, and non-parametric tests included the Kruskal-Wallis test and Wilcoxon signed-rank test. All statistical tests were two-tailed, and a Bonferroni correction was applied in all pairwise comparisons. Significance was considered at *p* < 0.05.

## 3. Results

### 3.1. Optimisation of material parameters

For each patient, a different end-diastolic pressure-volume relationship was obtained using the Klotz method [37] and used in the optimisation of the passive material parameters. The optimisation resulted in *in silico* end-diastolic volumes that closely matched the target *in vivo* values (Table 3). However, the Klotz method underestimated the unloaded LV volume (*V*_0K_) compared to the unloaded LV volume of the segmented geometry (*V*_0_) (Table 3). This discrepancy increased with an increase in *V*_0_.

**Table 3:** LV unloaded volumes, optimised material parameters, and the relative error in the end-diastolic volume and the mean squared error (MSE) of the LV end-diastolic pressure values obtained from comparisons between the *in silico* model and the target patient’s Klotz curve. *V*_0K_: Unloaded left ventricular volume determined with the Klotz method, *V*_0_: unloaded left ventricular volume based on ventricular geometry, *A*: scalar multiplier and *B*: exponential multiplier from initial optimisation algorithm, *A*_mod_: scalar multiplier and B_mod_: exponential multiplier from modified optimisation algorithm, and MSE: Mean squared error for LV pressure (Klotz curve versus *in silico* curve).

| Patient | $V_0$<br>(ml) | $V_{0K}$<br>(ml) | $A$ | $B$ | $A_{mod}$ | $B_{mod}$ | EDV Error<br>(%) | MSE<br>(mmHg) | $T_{max}$<br>(kPa) |
| --- | --- | --- | --- | --- | --- | --- | --- | --- | --- |
| R45A | 60 | 59 | 0.1000 | 1.8485 | - | - | 1.7857 | 1.0019 | 154 |
| N39A | 82 | 62 | 1.2270 | 1.5159 | - | - | 0.7407 | 0.3392 | 135 |
| R20A | 134 | 87 | 9.7529 | 0.3109 | 6.2000 | 1.6822 | 0 | 3.5994 | 200 |
| N18A | 152 | 87 | 28.8488 | 0.1000 | 14.2239 | 1.6822 | 0 | 6.5459 | 233 |
| N12A | 203 | 116 | 28.8500 | 0.1000 | 21.015 | 1.6822 | 0 | 15.1984 | 187 |
| N06D | 174 | 85 | 133.1495 | 0.1000 | 59.5059 | 1.6822 | 0 | 69.6533 | 110 |

The optimisation yielded substantially higher scalars *A* > 9 and lower exponential multipliers *B* < 0.4 than for models R45A and N39A, which resulted in linear *in silico* end-diastolic pressure-volume curves for patients R20A, N18A, N12A and N06D. However, Klotz [37] reported that end-diastolic pressure-volume relationships have a non-linear convex shape in common, irrespective of the species or heart condition. To improve models R20A, N18A, N12A and N06D, the optimisation algorithm for these four models was revised by (i) assigning *B* = 1.6822 as the average value from models R45A and N39A and (ii) changing the error function to the squared difference between the predicted and the patient-specific end-diastolic volume. This produced lower values for *A* for all four models (Table 3) and more non-linear end-diastolic pressure-volume curves, particularly for R20A and N18A. The accuracy of the passive material parameter optimisation was quantified for each model with the error of end-diastolic LV volume and the mean squared error of the LV pressure over the passive filling period (Table 3).

The final LV end-diastolic pressure-volume curves for each model after optimisation of the material parameters are displayed in Figure 2 (a). To compare the relative increase during the filling phase, the volume was normalised to that of the unloaded state, V_0_, using:

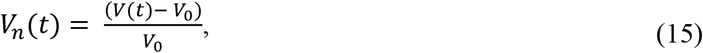

where V(t) is the non-normalised and V_n_(t) the normalised volume of the LV at each time point during the passive filling. Qualitatively, the slope and linearity of the curves increased with a decrease in the volume entering the LV during passive filling (i.e., EDV-ESV). Models R45A and N39A exhibited more compliant LV than R20A, N18A, N12A, and N06D, i.e. LV compliance decreased with a decrease in ejection fraction.

**Figure 2:**
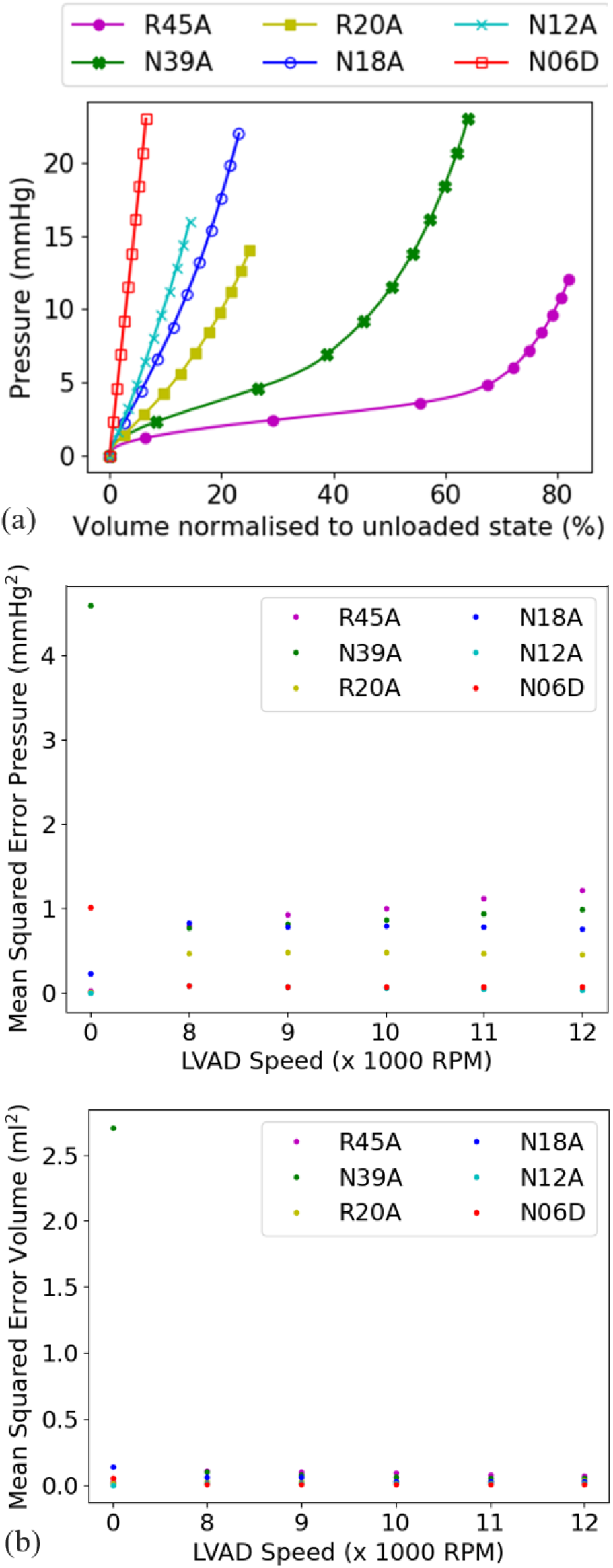
a) Calibrated *in silico* LV passive filling curves for each patient with volume normalised to the unloaded volume V_0_ (Table 3). Compliance of the LV myocardium increased with an increase in ejection fraction (represented by the numerical value in the model code). The model for patient N06D with the lowest ejection fraction of 6% had the stiffest LV myocardium. b) Solution convergence of the patient-specific models for multiple cardiac cycles expressed as mean squared error for LV pressure (top) and volume (bottom) of the last two cardiac cycles versus LVAD speed. The mean squared error is based on all pressure and volume values from the LV pressure-volume loops of the 5^th^ and 6^th^ cardiac cycle for models without LVAD support (LVAD speed of 0 RPM) and the 9^th^ and 10^th^ cardiac cycle for models with LVAD support.

The optimisation of the active material model yielded an average *T*_*max*_ of 170 ± 45 kPa, ranging from 110 kPa for N06D to 233 kPa for N39A (Table 3).

The optimised models reached solution convergence at the 6^th^ and 10^th^ cardiac cycle without and with LVAD support, respectively, based on the difference of LV pressure and volume between the respective last two cardiac cycles (Figure 2 b). All except one model achieved a mean square error of less than 1.3 mmHg^2^ for LV pressure and less than 0.2 ml^2^ for LV volume. The model without LVAD support for patient N39A exhibited mean square errors of 4.6 mmHg^2^ for pressure and 2.7 ml^2^ for volume.

### 3.2. Cardiac function

#### 3.2.1. Ventricular volumes and pressures

Models without LVAD accurately matched the *in vivo* LVEF and volume (from the patient’s clinical records) (Table 4 and Figure 3 a). For each patient, the target end-systolic *in vivo* volume was the *V*_0_ after segmentation (Table 4), and the end-diastolic volume was derived using clinical EF and *V*_0_. However, the predicted LV end-diastolic pressure deviated from the *in vivo* value by 21.2% ± 5.4%, −10.1% ± 2.1%, and 2.0% ± 15.6% for patients with an LVEF > 20%, LVEF < 20%, and all patients, respectively (Figure 3 b).

**Table 4:** Functional and dimensional parameters of LV and RV. Unloaded ventricular volume *V*_0_ of reconstructed cardiac geometries, end-diastolic volume (EDV) calculated using clinically recorded ejection fraction (*in vivo* EF) and *V*_0_, and ejection fraction predicted with finite element models (*in silico* EF).

| Patient | LV $V_0$<br>(ml) | LV EDV<br>(ml) | LVEF<br>*in vivo*<br>(%) | LVEF<br>*in silico*<br>(%) | RV $V_0$<br>(ml) | RV EDV<br>(ml) | RVEF<br>*in vivo*<br>(%) | RVEF<br>*in silico*<br>(%) |
| --- | --- | --- | --- | --- | --- | --- | --- | --- |
| R45A | 60 | 110 | 45 | 45.0 | 64 | 123 | 48 | 27 |
| N39A | 82 | 135 | 39 | 39.0 | 42 | 102 | 59 | 48 |
| R20A | 134 | 168 | 20 | 20.0 | 88 | 111 | 21 | 26 |
| N18A | 152 | 185 | 18 | 17.0 | 79 | 99 | 20 | 27 |
| N12A | 203 | 231 | 12 | 12.0 | 150 | - | - | 15 |
| N06D | 174 | 110 | 6 | 6.0 | 104 | 113 | 8 | 8 |

**Figure 3:**
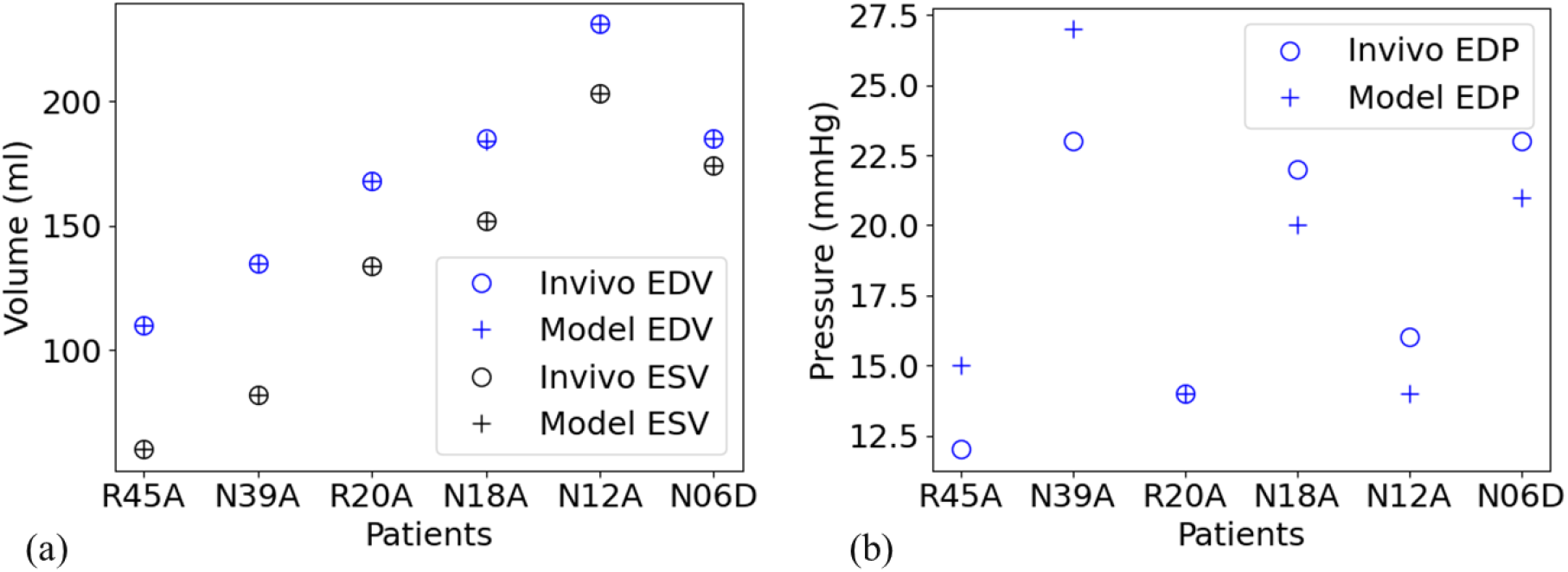
Comparison of *in vivo* and FE-predicted (Model) values for (a) LV volume at end-diastole (EDV) and end-systole (ESV) and (b) LV pressure at end-diastole (EDP). The models accurately replicated *in vivo* end-diastolic and end-systolic LV volumes but overestimated end-diastolic LV pressure for patients with EF > 20% and underestimated the LV pressure for patients with EF < 20%.

In models with LVAD support, the LV end-diastolic volume was lower than that without LVAD support (Student’s paired t-test, *p* ≥ 0.124). The 8,000 rpm LVAD yielded the lowest reduction in average LV end-diastolic volume of −9.0 ± 6.9 ml (n = 6, *p* = 0.141), whereas the 12,000 rpm LVAD yielded the highest reduction of −15.8 ± 13.8 ml for all models (*p* = 0.190).

The average RV end-diastolic volume of models with LVAD support increased marginally compared to those without LVAD support (*p* ≥ 0.0758, n = 6, Student’s paired t-test), except for patient R45A, who had slight increases of −0.8% at higher speeds of 11,000 and 12,000 rpm.

Compared to models without LVAD support for all patients, an increasing reduction in the average LV end-diastolic pressure was observed for increasing LVAD speeds, namely from −5.5 ± 3.4 mmHg (*p* = 0.0531) at 8,000 rpm to −8.7 ± 3.4 mmHg (*p* = 0.00819) at 12,000 rpm (n = 6, Student’s paired t-test).

The average RV end-diastolic pressure for all models increased slightly with the 8,000 rpm LVAD support (2.0 ± 0.5166 mmHg, *p =* 0.300) compared to no LVAD support. There was no change for the 10,000 rpm; however, there was a slight decrease for models with LVAD speeds of 9,000, 11,000 and 12,000 rpm (−0.7 ± 0.5164 mmHg, *p <* 4.165).

#### 3.2.2. Pressure-volume loops

The LV pressure-volume loops became triangular with LVAD compared to no LVAD, i.e. do not feature isovolumetric phases, and triangularity increases with increasing LVAD speed (Figure 4 a). The LV pressure-volume loops of models with low ejection fraction (N18A, N12A and N06A) shifted more downwards and to the left than those of models with higher ejection fraction (R45A and N39A). There was a noticeable rightward shift of the RV end-diastolic volume for the non-recovered patients (N39A, N18A, N12A and N06D), whereas marginal changes were observed for the recovered patients (R45A and R20A). Of note are the RV pressure-volume loops for model R45A, which were further to the right with higher end-diastolic volumes than the LV curves of 125 (IQR 0.75) ml versus 110 (IQR 0.75) ml, respectively, (*p* = 0.031, n = 6, Wilcoxon signed-rank test).

**Figure 4:**
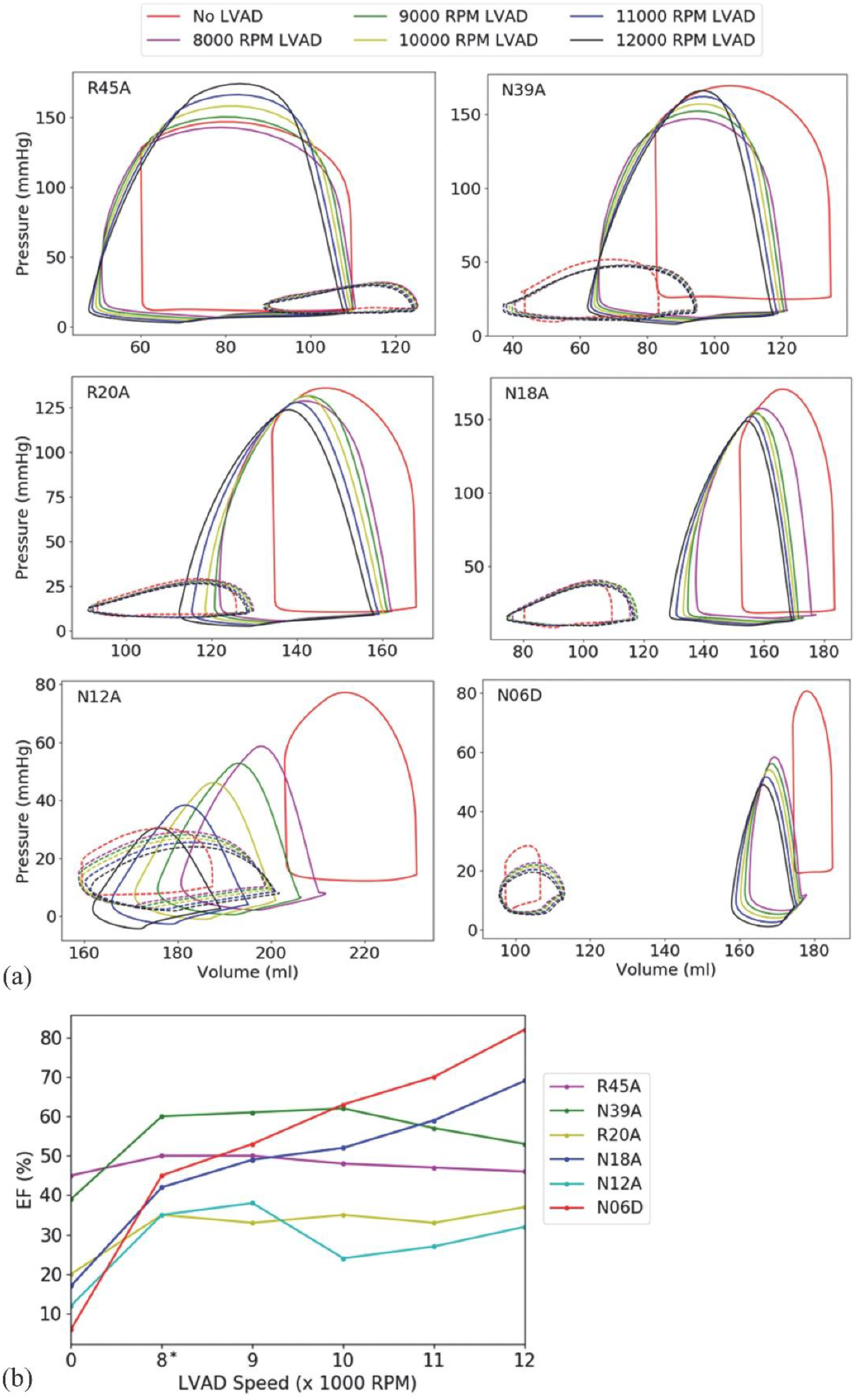
a) Pressure-volume loops for the LV (solid lines) and RV (dashed lines) versus LVAD speed for all patients. Apart from R45A, the LV pressure-volume curve shifted to the left and downwards, whereas the RV pressure-volume curve shifted to the right after implementing the LVAD support. b) Changes in the LV ejection fraction (EF) with an increase in LVAD speeds. The zero revolutions per minute (RPM) on the horizontal axis represents models without the LVAD. Increasing the LVAD speeds led to a steady increase in EF in some models, while there was a decrease or marginal change in EF in others. (* Indicates significant differences between the average ejection fraction for models without and with LVAD at each speed, *p* < 0.05).

#### 3.2.3. Ejection fraction

Implementing the LVAD in the models resulted in the LVEF (i) steadily increasing (N18A and N06D), (ii) fluctuating (R20A and N12A), (iii) reaching a maximum at an intermittent LVAD speed (N39A), and (iv) steadily decreasing (R45A) with LVAD speeds increasing from 8,000 to 12,000 rpm, see Figure 4 b. For the LVAD with 8,000 rpm compared to no LVAD support, the average LVEF improved by 21.3 ± 11.3 per cent points for all models (*p* = 0.0283, n = 6, Student’s paired t-test), see Table 4. For LVAD speeds above 8,000 rpm, there were slight increases in the average LVEF (*p* ≥ 0.0505) for all patients.

Notably, PPCM patients had a lower average EF in the LV than the RV without LVAD, whereas the average EF was higher in the LV than the RV with LVAD support, irrespective of the LVAD speed.

The average LV cardiac output improved with LVAD compared to no LVAD for all patients (2.7 ± 1.2 L/min) and ranged from 5.3 ± 0.9 L/min to 6.3 ± 3.0 L/min for 8,000 to 12,000 rpm LVAD speed (*p* > 0.0156, n = 6, Student’s paired t-test).

#### 3.2.4. LVAD contribution to stroke volume

The average LV stroke volume increased with LVAD compared to no LVAD (Figure 5, *p* > 0.0819, n = 6, Student’s paired t-test). For the 8,000 rpm LVAD compared to no LVAD, the increase in LV stroke volume increased with decreasing EF of the patients. The lowest and highest increase was observed for patients R45A (10%) and N06D (655%).

**Figure 5:**
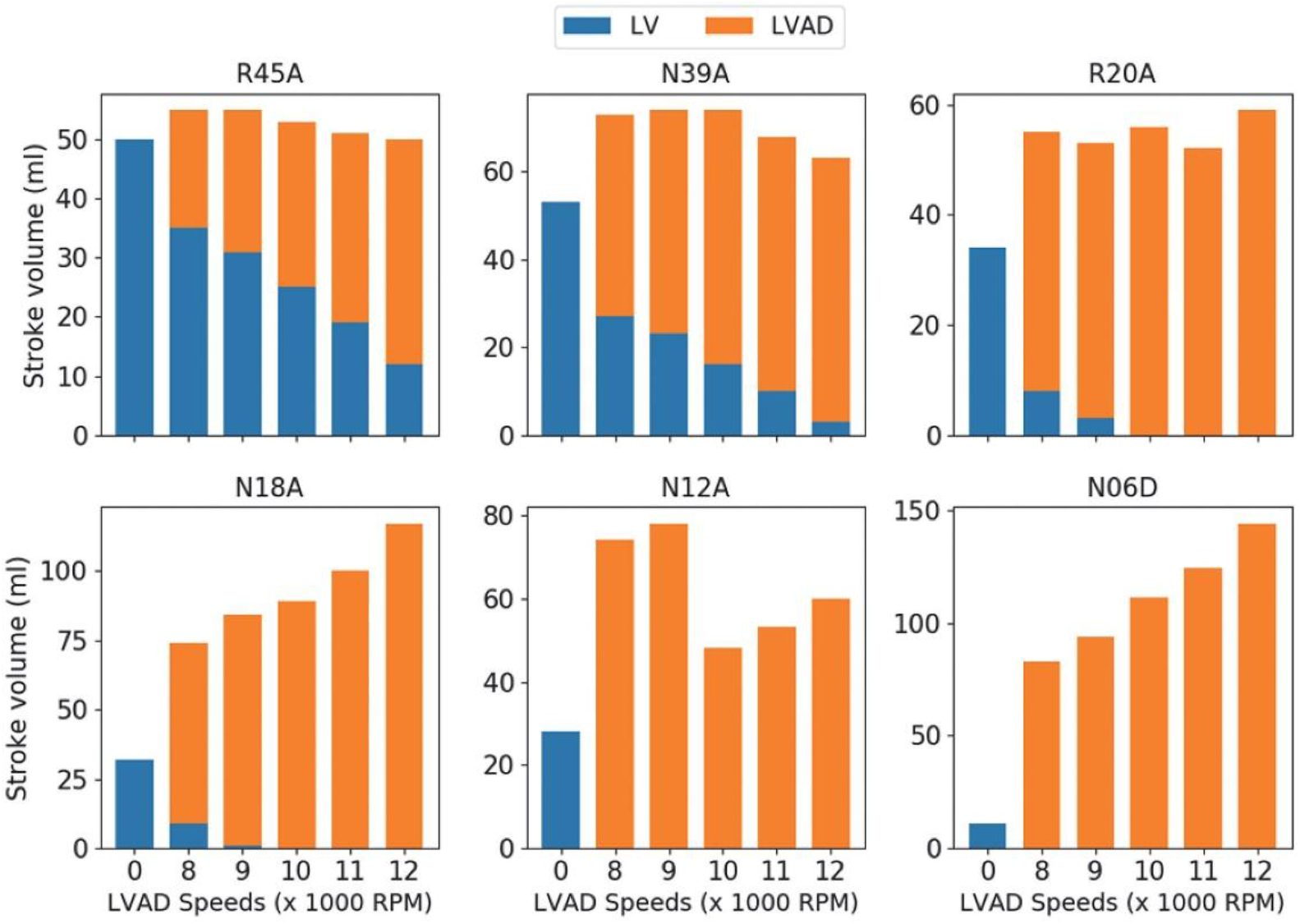
Variation of LV stroke volume contributions with LVAD speed for each PPCM model (zero revolutions per minute (RPM) represents models without LVAD). The LV and LVAD jointly contributed to stroke volume in the four models with EF = 18% and higher, while the LVAD was the only contributor in the two models with the lowest EF of 12% and 6%.

The LV and the LVAD jointly contributed to the stroke volume in models with higher EF (R45A, N39A, R20A and N18A), whereas the LVAD was the only contributor in the models with the two lowest EF (N12A and N06D). For joint contribution to stroke volume, the LV contribution decreased with a decrease in EF amongst models and with an increase in LVAD speed for each model.

#### 3.3. Myofibre stress

#### 3.3.1. Individual patients

The LV myofibre stress distribution (i.e. probability density curve) was different for models without LVAD support than for models with LVAD support, and it did not change with LVAD speed, except for patient R45A (Figure 6). For this patient, the stress distribution with and without LVAD was similar at end-diastole. The peak probability density was higher with LVAD support than without, and increased with increasing LVAD speed, for all patients.

**Figure 6:**
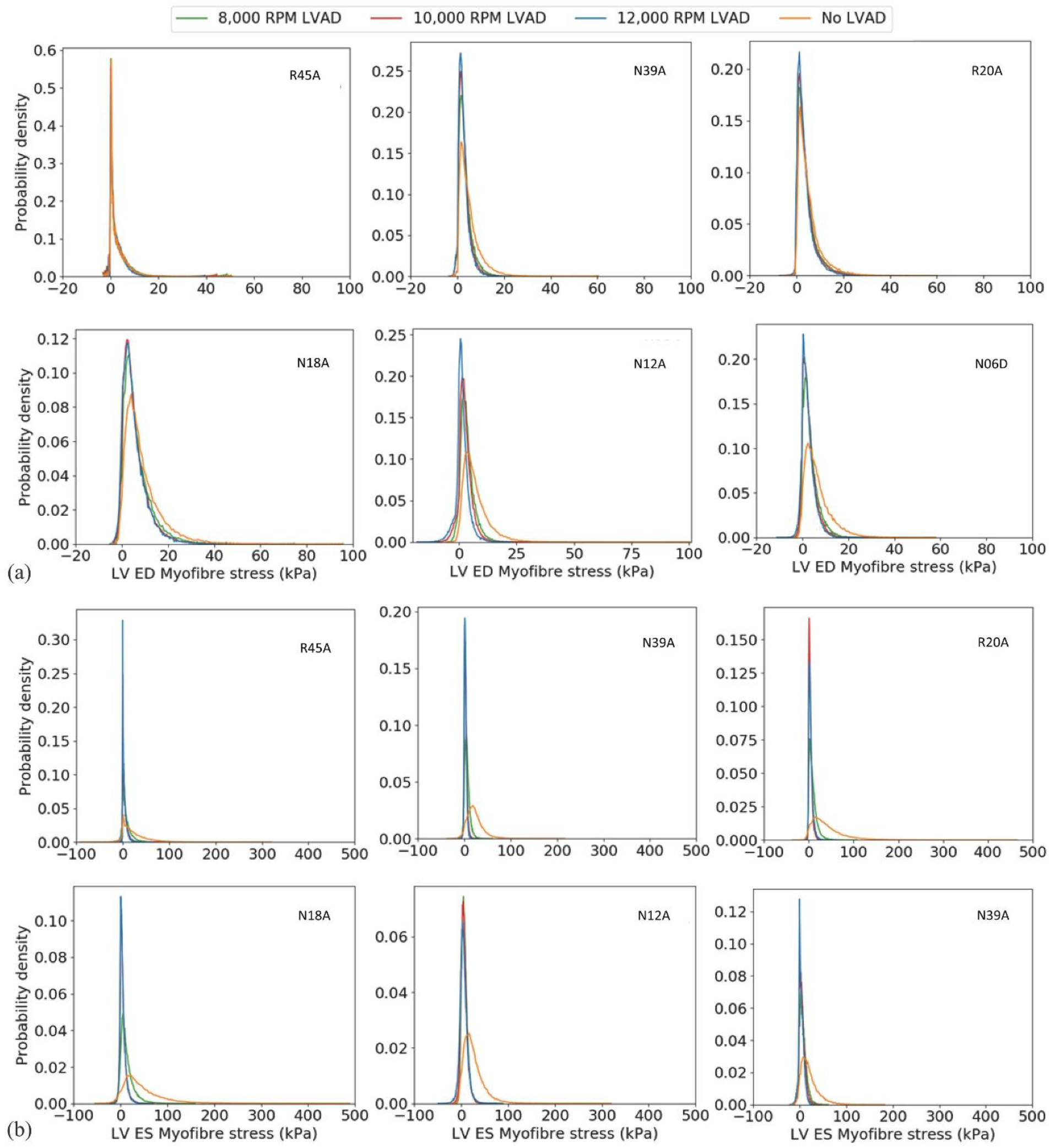
Probability density curves showing the LV myofibre stress distribution for each patient at (a) end-diastole (ED) and (b) end-systole (ES). LV myofibre stress distribution was different for models without LVAD support but similar for different LVAD speeds for each patient except R45A. For that patient, similar stress distributions were predicted with and without LVAD at end-diastole. The peak probability density was higher with than without LVAD support and increased with increasing LVAD speed.

The two main observations from the spatial distribution of myofibre stress (see representative plots for R45A and N06D in Figure 7) were:

**Figure 7:**
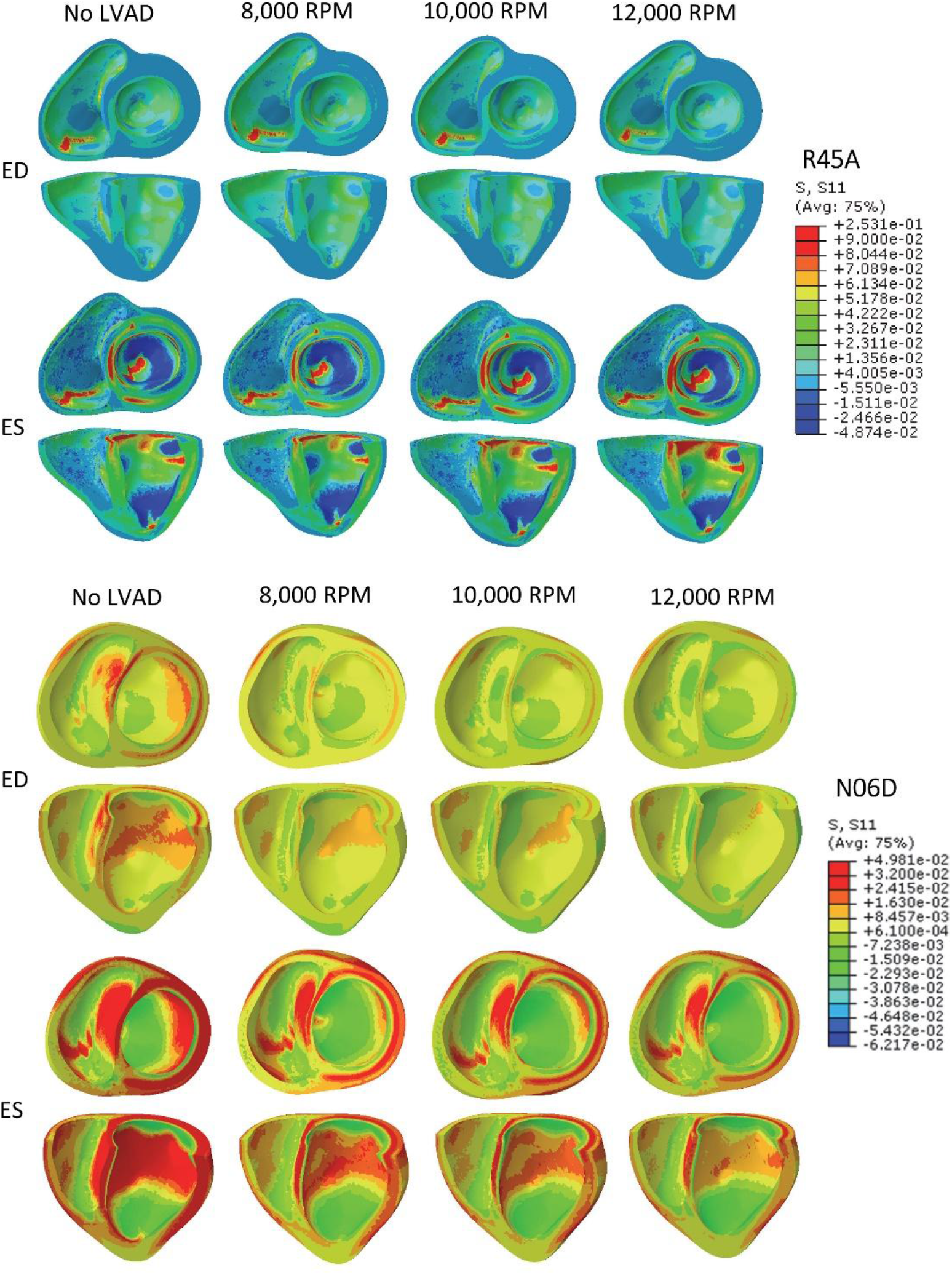
Representative contour plots illustrating two main observations for the biventricular myofibre stress in PPCM with increasing LVAD speed at end-diastole (ED) and end-systole (ES). Model R45A illustrates the first observation of a decrease in myofibre stress with an increase in LVAD speed at end-diastole, but an increase in myofibre stress with an increase in LVAD speed from 8,000 to 12,000 rpm at end-systole. This observation applies to models R45A and N39A. Model N06D illustrates the second observation with a decrease in myofibre stress with increasing LVAD speed, both at end-diastole and end-systole, which applies to models R20A, N18A, N12A and N06D.

- The end-diastolic myofibre stress decreased from no LVAD to LVAD support at 8,000 rpm and decreased from 8,000 to 12,000 rpm LVAD speed, whereas the end-systolic myofibre stress decreased from no LVAD to LVAD at 8,000 rpm but increased from 8,000 rpm to 12,000 rpm LVAD speed. This was observed for models R45A and N39A.
- For models R20A, N18A, N12A and N06D, the end-diastolic and end-systolic myofibre stress decreased from no LVAD to LVAD at 8,000 rpm and further decreased from 8,000 rpm to 12,000 rpm. This decrease in myofibre stress with increasing LVAD speed was smaller for R20A and N18A than for N12A and N06D.

#### 3.2.2. All patients

The patient’s mean myofibre stress in LV and RV was lower with LVAD than without LVAD for each patient, except for the RV end-diastolic myofibre stress that exhibited only marginal changes (Figure 8).

**Figure 8:**
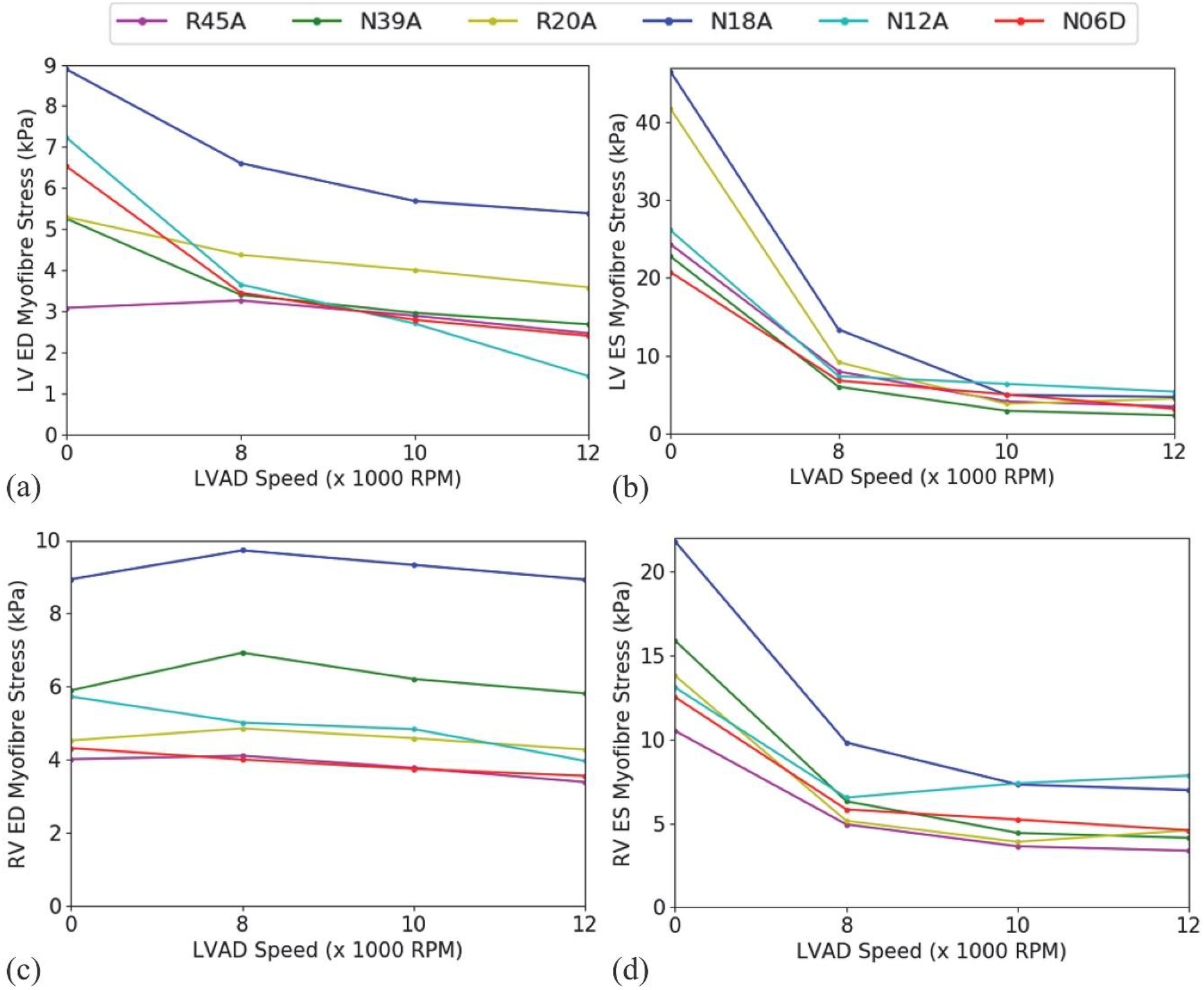
Patient mean myofibre stresses versus LVAD speed in LV and RV at end-diastole (ED) and end-systole (ES). The LVAD support led to a decrease in myofibre stress in the LV at end-diastole (a) and end-systole (b). In the RV, LVAD support decreased myofibre stress only at end-systole (d) and not at end-diastole (c). Zero revolution per minute (RPM) represent models without the LVAD.

In the LV at end-systole, the average myofibre stress was significantly lower with LVAD than without LVAD (between 3.6-fold decrease at 8,000 rpm and 7.7-fold decrease at 12,000 rpm, *p* < 0.00661, n = 6, Student’s paired t-test), whereas the decrease was smaller at end-diastole (between 1.4-fold at 8,000 rpm and 2-fold at 12,000 rpm, *p* > 0.938, n = 6, Wilcoxon signed-rank test) (Table 6).

**Table 5:** Average LV and RV ejection fraction (EF) without LVAD and with LVAD at different speeds for all patients (n = 6). * Represents statistical significance with p < 0.05 for pairwise comparisons with models without LVAD at each LVAD speed, including Bonferroni correction.

| LVAD Speed<br>(x1000 rpm) | LV EF<br>(%) | RV EF<br>(%) |
| --- | --- | --- |
| None | $23.2 \pm 15.5$ | $25.2 \pm 13.6$ |
| 8 | $44.5 \pm 9.57^*$ | $29.3 \pm 14.7$ |
| 9 | $47.3 \pm 10.2$ | $30.0 \pm 15.0$ |
| 10 | $47.3 \pm 15.4$ | $31.3 \pm 15.3$ |
| 11 | $48.8 \pm 16.4$ | $31.2 \pm 15.9$ |
| 12 | $53.2 \pm 19.2$ | $31.2 \pm 16.4$ |

**Table 6:** Average LV and RV myofibre stress for all patients at end-diastole and end-systole at different LVAD speeds. *Statistical significance for comparison with models without LVAD (*p* ≤ 0.01).

| LVAD Speed<br>(x1000 rpm) | End-diastolic stress |  | End-systolic stress |  |
| --- | --- | --- | --- | --- |
|  | LV (kPa) | RV (kPa) | LV (kPa) | RV (kPa) |
| 0 | 6.05 ± 1.98 | 5.57 ± 1.82 | 30.35 ± 10.90 | 14.62 ± 3.94 |
| 8 | 4.13 ± 1.28 | 5.78 ± 2.20 | 8.47 ± 2.63* | 6.43 ± 1.77* |
| 10 | 3.51 ± 1.17 | 5.42 ± 2.12 | 4.57 ± 1.20* | 5.32 ± 1.67* |
| 12 | 3.00 ± 1.36 | 4.99 ± 2.12 | 3.95 ± 1.12* | 5.26 ± 1.76* |

Similar to the observation for the LV, RV average myofibre stress was predicted to be significantly lower for models with LVAD than without LVAD at end-systole (between 2.3-fold decrease at 8,000 rpm and 2.8-fold decrease at 12,000 rpm, *p* ≤ 0.00348, n = 6, Student’s paired t-test). At end-diastole, the RV average myofibre stress was not significantly different with and without LVAD support (no change at 8,000 rpm and a 1.1-fold decrease at 12,000 rpm, *p* ≥ 1.31, n = 6, Student’s paired t-test).

### 3.4. Myofibre strain

#### 3.4.1. Individual patients

As observed for myofibre stress, the LV myofibre strain probability density was different without and with LVAD, but similar for different LVAD speeds at end-diastole and end-systole, except for patient R45A at end-diastole, with similar strain probability curves with and without LVAD.

Two trends were observed from the spatial distribution of biventricular myofibre strain (Figure 9):

**Figure 9:**
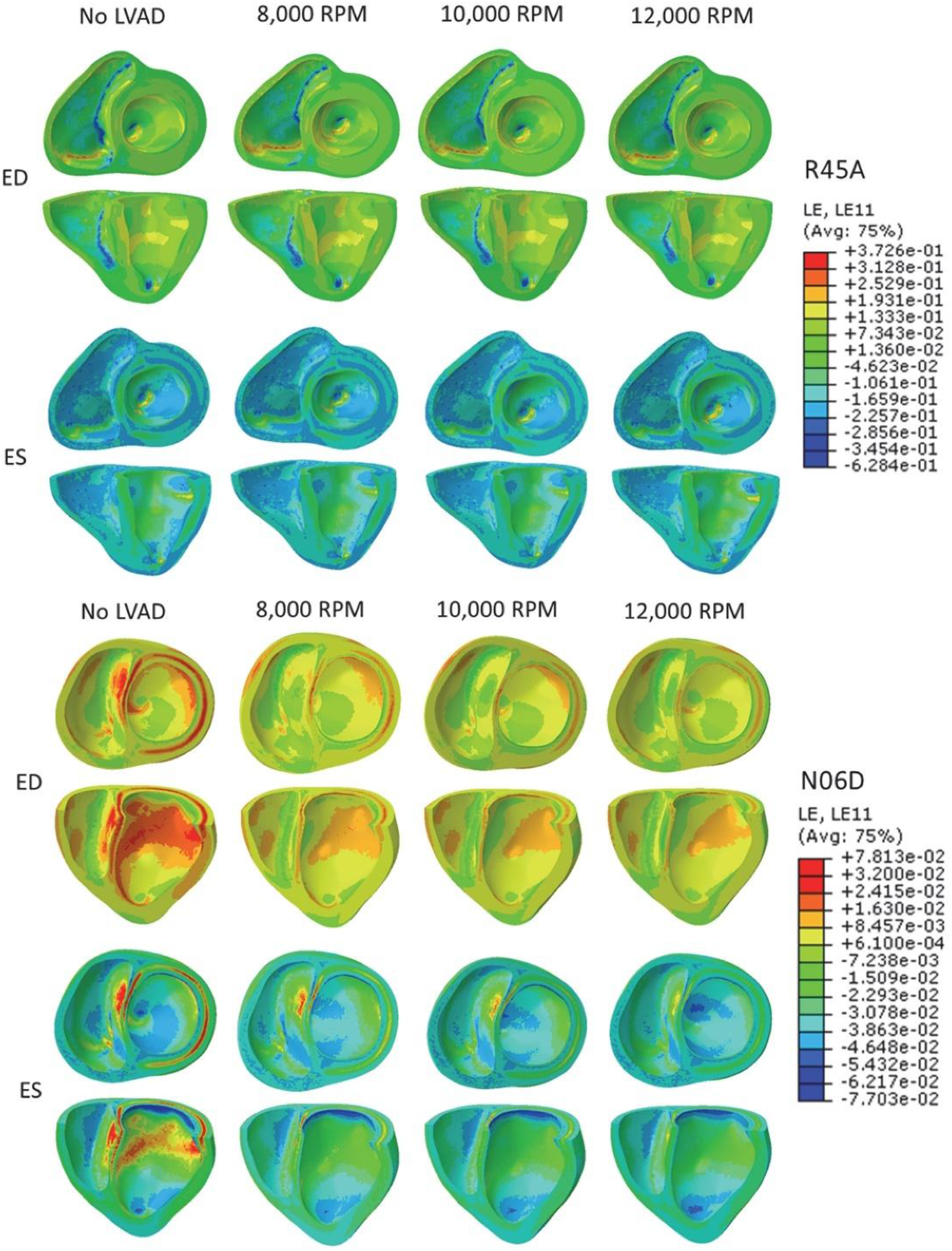
Representative contour plots displaying the two main trends of the biventricular myofibre strain distribution for the PPCM patients with and without the LVAD support at end-diastole (ED) and end-systole (ES). For patient R45A, end-diastolic myofibre strains decreased from no LVAD to the LVAD at 8,000 rpm and decreased further from 8,000 to 12,000 rpm LVAD speed. At end-systole, the myofibre strains decreased from no LVAD to the LVAD at 8,000 rpm but increased marginally with increasing LVAD speed from 8,000 to 12,000 rpm. For patient N06D, representative of all other patients, end-diastolic and end-systolic myofibre strain decreased from no LVAD to the LVAD at 8,000 rpm and decreased further from 8,000 to 12,000 rpm LVAD speed.

- The end-diastolic myofibre strain decreased from no LVAD to LVAD support at 8,000 rpm and decreased from 8,000 to 12,000 rpm LVAD speed, whereas the end-systolic myofibre strain decreased from no LVAD to LVAD support at 8,000 rpm but increased marginally with increasing LVAD speed from 8,000 to 12,000 rpm. This was observed for patient R45A.
- For the other five patients, the end-diastolic and end-systolic myofibre strain decreased from no LVAD to the LVAD at 8,000 rpm and decreased from 8,000 to 12,000 rpm LVAD speed.

#### 3.4.2. All patients

Marginal changes in the patient mean myofibre strain in LV and RV were observed with LVAD support compared to no LVAD. In the LV, the average myofibre strain was slightly lower with LVAD support than without the LVAD support at end-diastole and end-systole for the 8,000 rpm and 10,000 rpm LVAD (*p* ≥ 0.0764), whereas for the 12,000 rpm, there was a significant decrease at end-diastole (*p* = 0.0385) and a slight increase at end-systole (*p* = 2.543) (Table 7).

**Table 7:** Average LV and RV myofibre strain for all patients at end-diastole and end-systole for models without LVAD and with LVAD at different speeds. *Statistical significance for comparison with the model without LVAD (p < 0.05, n = 6, Student’s paired t-test).

| LVAD speed<br>(x1000 rpm) | End-diastolic strain |  | End-systolic strain |  |
| --- | --- | --- | --- | --- |
|  | LV (%) | RV (%) | LV (%) | RV (%) |
| None | $4.38 \pm 3.31$ | $2.37 \pm 2.19$ | $-6.68 \pm 3.51$ | $-8.94 \pm 4.30$ |
| 8 | $3.86 \pm 3.49$ | $2.41 \pm 2.23$ | $-6.52 \pm 3.44$ | $-7.19 \pm 3.85^*$ |
| 10 | $3.64 \pm 3.47$ | $2.33 \pm 2.17$ | $-6.30 \pm 3.04$ | $-6.60 \pm 3.38$ |
| 12 | $3.41 \pm 3.43^*$ | $2.26 \pm 2.13$ | $-6.76 \pm 3.00$ | $-6.98 \pm 3.34$ |

In the RV, the average myofibre strain marginally decreased with LVAD compared to no LVAD at end-diastole and end-systole for the 10,000 rpm and 12,000 rpm LVAD (*p* ≥ 0.0647), whereas for the 8,000 rpm, there was a slight increase at end-diastole (*p* = 2.5314) and a slight decrease at end-systole (*p* = 0.0293).

## 4. Discussion

This study investigated how disease severity affected the efficacy of partial LVAD support in South African PPCM patients using patient-specific finite element modelling to provide insights into therapeutic mechanisms for personalised LVAD treatment in PPCM.

### 4.1. Optimisation of material parameters

The mean passive material parameters for all PPCM patients, i.e. A = 17.05 ± 22.28 kPa and B = 1.68 ± 0.11, were substantially higher than those reported for the healthy model (A = 0.65 kPa and B = 1.00) [42] and heart failure model (A = 8.00 kPa and B = 4.00) [22]. This could imply that the LV myocardium is stiffer in PPCM than in the healthy case and in heart failure. The mean active tension parameter of the PPCM models (*T*_*max*_ = 170 ± 45 kPa) was higher than that for a healthy human heart (T_max_ = 143 ± 11.1 kPa) [43] but the same as the T_max_ = 170 kPa reported for heart failure in humans [22].

The method by Klotz [37] has been used to estimate a patient’s LV end-diastolic pressure-volume relationships, but it underestimated the initial volumes (V_0_) for PPCM patients (Table 3). Such discrepancies were observed in previous studies, for example, 2.92% [44], 38% [32] and 32% [22]. In another study, Dabiri [42] constrained the initial volume to the *V*_0K_ determined by the Klotz method. Patient N06D with the highest discrepancy between *V*_0_ and *V*_0K_ yielded the highest optimisation mean squared error (Table 3). Therefore, there is a need to improve the calibration of patient-specific passive material parameters for patients with similar characteristics.

Despite the limitations in the material parameter optimisation, the optimised patient-specific models ents produced cardiac function metrics similar to the target *in vivo* metrics (Table 4 and Figure 3). Models with LVAD required more cardiac cycles to achieve solution convergence than models without LVAD since the LVAD changed the conditions in the circulatory system, whose fluid exchanges are driven by active contraction and pressure differences between cavities.

### 4.2. Cardiac function

LVAD support decreased end-diastolic volumes and pressures in the LV for all models, but increased either the volume or pressure in the RV for some models, except R45A. These changes were amplified with increasing LVAD speed, indicated by the left and downward shift of the LV pressure-volume loops and the rightward shift of the RV pressure-volume loops, particularly for models with lower LVEF (N18A, N12A and N06A, Figure 4 a). The increasingly triangular shape of LV pressure-volume loops indicated continuous unloading of the LV by the LVAD during the isovolumetric phases of the cardiac cycle (Figure 4 a), consistent with findings by Sack [22].

While the LVAD provides haemodynamic unloading of the LV, the resultant increase in systemic blood flow increases the RV workload, left septal shift and RVEDV, altering RV geometry and function [22, 45]. For this reason, RV failure is often a contraindication to the eligibility of patients for LVAD support.

In the current study, there was an increase in RV end-diastolic volume due to LVAD support, which resulted in an increase in RV ejection fraction except for patient R45A. Interestingly, patient R45A experienced decreases in RV end-diastolic volumes (−0.8%) and pressures (−13%) at high LVAD speeds (11,000 - 12,000 rpm) compared to models without LVAD. This may indicate better RV compliance and capacity to adapt at higher LVAD speeds for models with higher ventricular ejection fraction at baseline (LVEF: 45% and RVEF: 48%, Table 4). *In silico* predictions could, therefore, potentially determine the likelihood of myocardial recovery with LVAD or drug therapy, in addition to determining patient-specific LVAD speeds that would be less likely to incur worsening RV failure.

The improvements in the LVEF due to LVAD treatment predicted for PPCM (between 21 ± 11.3 per cent points at 8,000 rpm and 30 ± 28.2 per cent points at 12,000 rpm) were greater than those reported for chronic heart failure (between 0.9 per cent points at 8,000 rpm and 6.5 per cent points at 12,000 rpm) [22]. For some patients, the improvement in LVEF reached a maximum at an intermediate LVAD speed and declined or fluctuated for further increasing speed (Figure 4 b). These findings suggest the existence of patient-specific and/or disease-specific optimal LVAD speeds and emphasise the importance of computational modelling to advance personalised LVAD treatment in patients with heart failure.

PPCM patients benefited functionally from LVAD treatment, with a larger EF predicted for the LV than the RV, with the LVAD support similar to the healthy case [30] compared to the lower EF in the LV than the RV in models without LVAD support (Table 5). This further illustrates the ability of the LVAD to provide more benefits for patients with lower LVEF than those with higher LVEF.

The lowest LVAD speed of 8,000 rpm was sufficient for all PPCM patients to achieve an LV cardiac output within the reference range of 3.3 - 7.3 L/min for healthy women [46], namely between 4.24 L/min for R45A (with the highest EF) and 6.40 L/min for N06D (with the lowest EF). However, only the highest LVAD speed of 12,000 rpm increased the average LVEF (53.2% ± 19.2%) close to the reference value of LVEF ≥ 54% for healthy women [30, 47].

The LV and LVAD co-contributed to the stroke volume for four patients with LVEF ≥ 18%. For these patients, the proportion of the LV contribution decreased with increasing LVAD speed, and the LVAD was the sole contributor to LV output at higher speeds from 10,000 rpm for patient N18A and 11,000 rpm for patient R20A with LVEF = 18% and 20%, respectively (Figure 5). For patients with the lowest LVEF of 6% and 12%, the LVAD solely provided the ventricular output at all speeds. These findings concur with a previous study that suggested that mechanical support from the LVAD at high speeds (12,000 rpm) excessively reduces LV volume and results in reduced cardiac output compared to lower LVAD speeds [22]. The relationship between LVAD speed and the work required by the patient’s ventricle may affect the functional recovery of the LV. Therefore, patient-specific selection of the LVAD speed and adjustment during the recovery process to allow for increasing ventricular function may offer the potential to achieve optimal support in PPCM.

### 4.3. Myocardial stress

LVAD support resulted in a substantial reduction in end-systolic LV myofibre stress from 30.4 ± 10.90 kPa to between 8.5 ± 2.6 kPa and 3.95 ± 1.1 kPa (for 8,000 and 12,000 rpm, respectively), i.e. lower than 9.3 ± 10.3 kPa [43] and 16.5 ± 4.7 kPa [48] in healthy humans. However, the reduction of end-diastolic LV myofibre stress in PPCM through LVAD support did not reach values for healthy humans of 0.3 ± 1.4 kPa [48] and 2.2 ± 0.6 kPa [43].

The LVAD reduced LV and RV myofibre stresses to lower levels in PPCM than in heart failure reported in a previous study [22]. The reduction in end-systolic LV myofibre stress by LVAD support was larger for PPCM (−72.0%) than for chronic heart failure (−6.7%) [22] despite similar myofibre stress without LVAD, i.e. 30.2 ± 31.5 kPa and 35.7 ± 5.6 kPa [22].

The chronic heart failure in Sack [22] was due to ischemia, which may explain the lower myofiber stress reduction compared to PPCM. PPCM is a cardiac muscle pathology whose primary insult (pregnancy) does not persist like in other heart failure conditions, e.g. due to ischemia (where part of the myocardium becomes non-functional and scarred) or genetic cardiomyopathies. Loyaga-Rendon [12] reported a higher survival rate for PPCM patients (83%) than non-ischemic cardiomyopathy (65%) and ischemic cardiomyopathy patients (64%), two years after receiving mechanical circulatory support. The LVAD (n = 1123) was the most used device compared to the bi-ventricular assist device (n = 121) and the total artificial heart (n = 14) [12]. Mechanical circulatory support is given to severe acute PPCM patients who do not respond to drug therapy alone [10]; however, reasons for the higher recovery and survival rates in PPCM than in other forms of heart failure are not well understood and need further investigation.

Long-term use of LVAD is associated with adverse events, including right ventricular failure (Hitzeroth et al. 2020, Rich et al. 2017). The current study predicted higher myofibre stress in the RV than the LV at each LVAD speed, yet myofibre stress was lower in the RV than the LV without LVAD (Table 6). Due to the interdependence between LV and RV, the effective unloading of the LV by the LVAD is reported to cause volume and pressure overload and elevated myofibre stress in the RV [22, 23]. Similar to a study on chronic heart failure [22], the current study with six PPCM patients with different LVEF observed ventricular interdependence but not worsening RV failure due to LVAD support, typically indicated by elevated end-diastolic RV pressure of > 15 mmHg [49, 50]. The increase in RV ejection fraction as a result of increased RV preload could indicate better RV compliance and capacity in PPCM to adapt to the additional volume from the LVAD.

### 4.4. Myocardial strain

Data on myofibre strains in the literature are scarce for humans, but more readily available for other species, such as porcine. For PPCM, the end-systolic average myofibre strain was significantly higher in the RV than the LV, similar to findings for heart failure in porcine by Sack [32].

LV mean myofibre strain in PPCM without LVAD (end-diastole: 4.4% ± 3.3%, end-systole: −6.7% ± 3.5%) was lower than for healthy hearts at both time points (end-diastole: 9.6% ± 6.7%, end-systole: −9.8% ± 5.0%) [32]. Compared to heart failure (end-diastole: 6.9% ± 4.3%, end-systole: −5.4% ± 6.5%) [32], PPCM myofibre strain was lower at end-diastole but similar at end-systole.

LVAD support reduced myofibre strain in PPCM (Table 7) to below that for healthy subjects (at end-diastole and end-systole) and heart failure (at end-diastole but not end-systole) [32]. The end-systolic LV myofibre strain was slightly higher in PPCM with LVAD support than in heart failure [32]. The continuous unloading during diastole could have restricted the full relaxation of the LV at end-diastole and thus the lower strains. At end-systole, the LVAD could have further reduced the LV volume and enhanced contraction, resulting in the higher strain.

### 4.5. Limitations

The small sample size of six PPCM patients and the unequal numbers of recovered (two) and non-recovered patients (four) limited generalised conclusions about PPCM and the impact of LVAD support in PPCM. The diversity in the cardiac conditions of the six patients aimed to represent PPCM of different severity. While the diversity and small sample size may have introduced bias or overfitting for a specific PPCM condition, the results of this study nevertheless provide insights into PPCM cardiac biomechanics and tailoring LVAD support to disease severity for optimal personalised efficacy. Future studies should involve larger patient numbers for the different PPCM conditions and equal group sizes to confirm the current results.

In the current study, the biventricular geometries truncated at the base of the ventricles did not include some anatomical structures, such as papillary muscles and atria. Future research may consider incorporating these anatomical features to advance cardiac computational models.

The lack of *in vivo* data, such as ventricular pressure and myocardial strain, restricted the validation of the PPCM models in this study. Nonetheless, the model calibration used has been validated [32] and produced reasonable results. This is confirmed in the current study as the PPCM myofibre stresses were similar to heart failure stresses, as may be expected, and elevated compared to myofibre stresses in the healthy heart. In the future, more PPCM models should be developed and validated using *in vivo* pressure and strain data.

The material law used in the PPCM models activates contraction at the same time everywhere in the ventricles compared to the more realistic propagation of the electrical activation from the sinoatrial node to the atrioventricular node, bundle of His and Purkinje fibres. For simplicity, the heart rate for all the models was 77 beats per minute. In the future, electrophysiology and the patient’s heart rate should be accounted for in the PPCM models.

The optimisation of patient-specific constitutive parameters was performed using LV measurements. This could explain the high discrepancies in the *in vivo* and *in silico* ejection fraction of the RV in the current study. Future studies should incorporate patient-specific RV quantities in the material parameter optimisation. Further, the different scales of the terms in Eqns. (7) and (8), respectively, the fixing of parameter B and the identical value for parameter B for four models may have introduced bias in the passive material optimisation. Separate normalisation for terms of different scales and fully automated optimisation regimes are suggested for future studies.

## 5. Conclusions

The current study demonstrated the potential of personalised treatment to improve partial LVAD support in PPCM patients with different disease severity in South Africa. The functional improvement was greater for PPCM patients with lower EF resulting from higher disease severity than for those with higher EF and lower disease severity. The results also suggest that partial LVAD support is more beneficial for patients with PPCM than those with chronic heart failure due to ischemia. The developed patient-specific models offer a tool for further research into the characterisation and treatment of PPCM. For example, identifying specific distinctive characteristics of PPCM may enable differential diagnosis of this condition, leading to more timely interventions and management. Focus of further research into LVAD support in PPCM should include patient- and disease-stage specific treatment options to guide clinical decisions and optimise personalised therapy outcomes.

## Abbreviations

cMRI: Cardiac magnetic resonance imaging
ED: End-diastole, end-diastolic
EDV: End-diastolic volume
EF: Ejection fraction
ES: End-systole, end-systolic
FE: Finite element
IQR: Inter-quartile range
LV: Left ventricle, left ventricular
LVAD: Left ventricular assist device
LVEF: Left ventricular ejection fraction
LVLS: Left ventricular long-axis shortening
PPCM: Peripartum cardiomyopathy
PCWP: Pulmonary capillary wedge pressure
rpm: Revolutions per minute
RV: Right ventricle, right ventricular
RVEF: Right ventricular ejection fraction
SV: Stroke volume

## Ethics statement

The protocol and procedures for the current study were approved by the Faculty of Health Sciences Human Research Ethics Committee of the University of Cape Town (HREC REF: 752/2018). The study conformed to the standards set by the latest version of the Declaration of Helsinki, except for registration in a database. This study only involved the use of existing observational data from the IMHOTEP clinical registry of cardiomyopathy patients, and no experiments were conducted on human participants. Data collection for the IMHOTEP registry was also approved by the Faculty of Health Sciences Human Research Ethics Committee of the University of Cape Town (HREC 766/2014). Data were only collected from participants who provided written informed consent, including for data reuse in different research studies beyond the life and specific remit of the ethics approval, such as the current study. The identification of patients has been anonymised.

## Data availability

Computational models and data supporting this study are available on the University of Cape Town’s institutional data repository ZivaHub under the DOI https://doi.org/10.25375/uct.26047783 as Nagawa J, Sack KL, Nchejane NJ, Motchon YD, Sirry MS, Kraus S, Davies NH, Ntusi NAB, Franz T. Software and data for Patient-specific in silico prediction of outcomes of partial continuous-flow LVAD treatment in peripartum cardiomyopathy. ZivaHub, 2024, DOI: 10.25375/uct.26047783.

## Acknowledgements

The authors thank Associate Professor Sulaiman Moosa, Head of the Division of Radiology, University of Cape Town, for helping with the data transfer during the study, and the Dassault Systèmes Living Heart Project for providing Abaqus licenses.

## Credit Author Contributions

Juliet Nagawa: Data curation, Formal analysis, Investigation, Methodology, Project administration, Software, Validation, Visualization, Writing – Original Draft Preparation, Writing – Review & Editing

Kevin L. Sack: Conceptualization, Methodology, Software, Supervision, Writing – Review & Editing

N. John Nchejane: Methodology, Writing – Review & Editing

Y. Dodzi Motchon: Methodology, Writing – Review & Editing

Mazin Sirry: Methodology, Software, Supervision, Writing – Review & Editing

Sarah Kraus: Conceptualization, Data curation, Resources, Writing – Review & Editing Neil H. Davies: Conceptualization, Supervision, Writing – Review & Editing

Ntobeko A.B. Ntusi: Conceptualization, Data curation, Resources, Writing – Review & Editing

Thomas Franz: Conceptualization, Data curation, Funding acquisition, Methodology, Project administration, Resources, Supervision, Validation, Writing – Original Draft Preparation, Writing – Review & Editing

